# Unique plasma metabolite signature for adolescents with Klinefelter syndrome reveals altered fatty acid metabolism

**DOI:** 10.1101/2021.08.30.21262854

**Authors:** Shanlee M Davis, Rhianna Urban, Angelo D’Alessandro, Julie Haines, Christine Chan, Megan Kelsey, Susan Howell, Nicole Tartaglia, Philip Zeitler, Peter Baker

## Abstract

Conditions related to cardiometabolic disease, including metabolic syndrome and type 2 diabetes, are common among men with Klinefelter syndrome (KS).The molecular mechanisms underlying this aberrant metabolism in KS are largely unknown, although there is an assumption that chronic testosterone deficiency plays a role. This cross-sectional study compared plasma metabolites in 31 pubertal adolescent males with KS to 32 controls of similar age (14 ± 2 yrs), pubertal stage, and body mass index z-score (0.1 ± 1.2), and then between testosterone treated (n=16) and untreated males with KS. The plasma metabolome in males with KS was distinctly different from controls, with 22% of measured metabolites having a differential abundance and seven metabolites nearly completely separating KS from controls (AUC>0.9, p<0.0001). Multiple saturated free fatty acids were higher in KS while mono- and polyunsaturated fatty acids were lower, and the top significantly enriched pathway was mitochondrial ß-oxidation of long-chain saturated fatty acids (enrichment ratio 16, p<0.0001). In contrast, there were no observed differences in metabolite concentrations between testosterone-treated and untreated individuals with KS. In conclusion, the plasma metabolome profile in adolescent males with KS is distinctly different from males without KS independent of age, obesity, pubertal development, or testosterone treatment status, and is suggestive of differences in mitochondrial ß-oxidation.

## Introduction

Klinefelter syndrome (KS), or 47,XXY, is strongly associated with a high risk for cardiometabolic comorbidities including dyslipidemia, type 2 diabetes, and cardiovascular disease.^1^ Historically, chronic untreated hypogonadism, a nearly universal finding in adult men with KS, has been implicated as the underlying pathologic mechanism leading to this increased risk.^2^ As an anabolic steroid, testosterone has effects on body composition and mitochondrial metabolism, which are widely accepted contributors to insulin resistance.^3^ The hypogonadal pathology assumption remains challenged given cardiometabolic disease risk factors are identified in younger cohorts of boys with KS naïve to years of hypogonadism, as well as findings that suggest testosterone replacement does little to improve cardiometabolic disease in adults.^4, 5^ Therefore, there is a need to deepen our understanding of the molecular mechanisms underlying the high cardiometabolic risk in KS.

Metabolomics, or the study of small molecules in biological samples, is a powerful tool to inform metabolic processes in disease states. Metabolomics is a comprehensive approach to assessing systems biology, encompassing both endogenous metabolic processes and exogenous exposures. It can be useful for disease diagnosis, prognosis, risk prediction, therapeutic development and assessment of response, and understanding pathophysiology, particularly when the underlying molecular mechanisms of a disease state are unknown.^6^ Of particular relevance to KS, metabolomics has been extensively used in characterizing cardiometabolic-related disease states, including metabolic syndrome and type 2 diabetes,^7^ as well as in the setting of male hypogonadism with testosterone-replacement therapy.^8^ Analysis of circulating levels of substrates and intermediate metabolites from intra-mitochondrial pathways, particularly ß- oxidation of fatty acids, offers a non-invasive assessment of mitochondrial metabolism – a viable strategy to predict intracellular metabolic states from extracellular metabolomics data.^9^ When fatty acid oxidation is impaired, as in the case with multiple cardiometabolic disorders, the reliance on protein catabolism to provide tricarboxcylic acid (TCA) cycle intermediates increases. As a result, the metabolite profile includes an increase in intermediates of fatty acid oxidation (classically long-chain acylcarnitine species) as well as branched-chain amino acids.^10^ These metabolites are suggested to not only be biomarkers of disrupted mitochondrial metabolism but also may contribute to progression of insulin resistance.

A few studies have investigated specific metabolites in KS, such as steroids, melatonin, and markers of bone metabolism,^11-13^ however a more global approach presents a unique opportunity to better understand aberrant metabolism in this population. The aim of this study was to compare the plasma metabolic signatures of adolescents with KS to controls, as well as between testosterone-treated and untreated males with KS. We hypothesized that we would observe a pattern suggestive of aberrant mitochondrial metabolism, and that testosterone would improve this metabolic profile.

## Methods

### Overall Study Design

This was a cross-sectional study of untargeted metabolite concentrations in the plasma of 31 males with KS compared to 33 controls selected for similar age, pubertal stage, and BMI.

### Study Participants and Procedures

Pubertal adolescents 12-17 years of age with KS were recruited to participate in a study on cardiometabolic health (COMIRB #16-0248). Prepubertal exam, hypertension, diabetes, or exercise restrictions were exclusionary. Additionally, boys that were being treated with testosterone had to be on treatment for at least a year. Participants with KS were recruited from the eXtraordinarY Kids Clinic and Research Program and advertisement through AXYS advocacy organization. All participants provided written assent and a parent provided written consent prior to any study procedures. All 31 participants had a fasting venous blood draw with blood collected in a sodium heparin tube. Tubes were centrifuged at 2000g for 15 minutes and plasma was aliquoted into cryovials and stored at -80° Celsius until batch analysis.

Plasma that had been obtained and processed in an identical manner from fasting healthy males participating in the Health Influences of Puberty (HIP) Study (NCT01775813) and the Glycemic Monitoring in Cystic Fibrosis (GEM) Study (NCT02211235) were used as a comparison group. Out of a pool of >100 participants in these studies with permission to use these samples for additional research, 37 pubertal males age 12-17 were selected based on age and BMI similar to the KS cohort. Of those, 33 had usable plasma and one was later excluded due to outliers for most metabolite concentrations, resulting in the analytic cohort in Table 1.

**Table 1.** Characteristics of the analytic sample

| Table 1. Characteristics of the analytic sample |  |  |  |
| --- | --- | --- | --- |
|  | KS (n=31) | Controls (n=32) | p-value |
| Age (yrs) | 14.5 ± 1.7 | 14.8 ± 1.7 | 0.49 |
| BMI z-score | 0.24 ± 1.51 | -0.05 ± 0.93 | 0.36 |
| Pubertal stage |  |  | 0.14 |
| Tanner 2 | 2 (6.5%) | 4 (12.5%) |  |
| Tanner 3 | 0 (0%) | 4 (12.5%) |  |
| Tanner 4 | 17 (54.8%) | 16 (50%) |  |
| Tanner 5 | 12 (38.7%) | 8 (25%) |  |
| Testosterone treatment | 16 (52%) | n/a | n/a |

### Metabolomics analysis

Plasma samples were analyzed in the University of Colorado School of Medicine Metabolomics Core as previously described.^14^ In brief, plasma metabolites were extracted with a solution of methanol, acetonitrile, and water (5:3:2 *v/v/v*) and 10 µl were injected for analysis via ultra-high-pressure liquid chromatography coupled to mass spectrometry on a Thermo Vanquish UHPLC – Thermo Q Exactive MS system. Compound identification was based on intact mass, ^13^C isotope pattern, and retention times from an in-house reference of over 5000 analytical standards. Relative metabolite abundance for each sample was reported. Analytical details are extensively provided in technical notes^15^ and previous applications to similar matrices.^16, 17^

### Statistical analysis

Data analysis was performed in MetaboAnalyst 5.0. Data were normalized by sum and auto-scaled (mean-centered and divided by the standard deviation of each variable). Metabolite abundance was compared between KS and control groups (followed by testosterone treatment status) with fold change and unpaired Mann Whitney tests. Adjustment for multiple comparisons was performed with the two-stage step-up method of Bejamini, Krieger and Yekutieli for calculation of the false discovery rate (FDR).^18^ Significance was set at q≤0.05 for definitive group differences, although comparisons resulting in q<0.15 were included in exploratory pathway analyses for hypothesis-generating integration of the results. For visualization, GraphPad Prism v9.0.2 was used to generate a volcano plot identifying metabolites with a fold change >1.5 and FDR <0.05. To determine if a unique metabolome profile is present in KS, Partial Least Squares – Discriminant Analysis (PLS-DA), a variant of principal component analysis that is considered the gold standard for binary classification of metabolomics datasets, was conducted.^19^ Receiver operating characteristic (ROC) curve analysis was conducted to identify potential biomarkers of KS, defined as metabolites with an area under the curve (AUC) >0.9. Finally, to help interpret the data in respect to normal human metabolism, we referenced the small molecule pathway database (SMPD) metabolite set library to determine enrichment for metabolic pathways, with FDR threshold of 5%.

## Results

### KS vs Controls

A total of 157 metabolites were identified in the samples using a targeted data analysis focused on metabolites central to energy and redox metabolism. Of these, 35 (22%) differed in abundance between KS and controls with an FDR <0.05 (Figure 1a). Most of these were fatty acids, with nearly all saturated fatty acids higher in KS and mono- and polyunsaturated fatty acids lower (Figure 1b). PLS-DA revealed a distinct signature for KS (Figure 1c), with the highest predictive ability inclusive of the first three components (accuracy 0.97, R2=90, Q2=0.84). Several metabolites were identified as potential biomarkers of KS with an AUC of >0.9 (Figure 1d) including lysophosphatidylcholine (AUC 1.0), the medium, odd-chain fatty acids undecanoic acid (AUC 1.0) and nonanoic acid (AUC 0.99), the long, even-chain fatty acids stearic acid (AUC 0.97) and palmitic acid (AUC 0.97), and prostaglandin E2 (AUC 0.90). A number of SMPD metabolic sets (31/99) were significantly enriched with an FDR up to 5%, with the top 25 shown in Figure 2. Mitochondrial ß-oxidation of long chain saturated fatty acids was the top enriched pathway with both a high enrichment ratio and strong significance. Several other enriched pathways also support aberrant mitochondrial metabolism, including fatty acid metabolism, oxidation of branched chain fatty acids, ß-oxidation of short chain saturated fatty acids, linoleic acid metabolism, and propanoate metabolism. Additional pathways of interest include plasmalogen synthesis and ß-oxidation of very long chain fatty acids, which are both part of peroxisomal metabolism that is closely tied with mitochondrial metabolism. There were also several amino acid metabolism pathways enriched in pathway analysis, including arginine/proline, urea cycle, tyrosine, aspartate, and propionate metabolism. Finally, there was enrichment of several steroid hormone pathways, including androgen and estrogen, androstenedione, and estrone metabolism.

**Figure 1.**
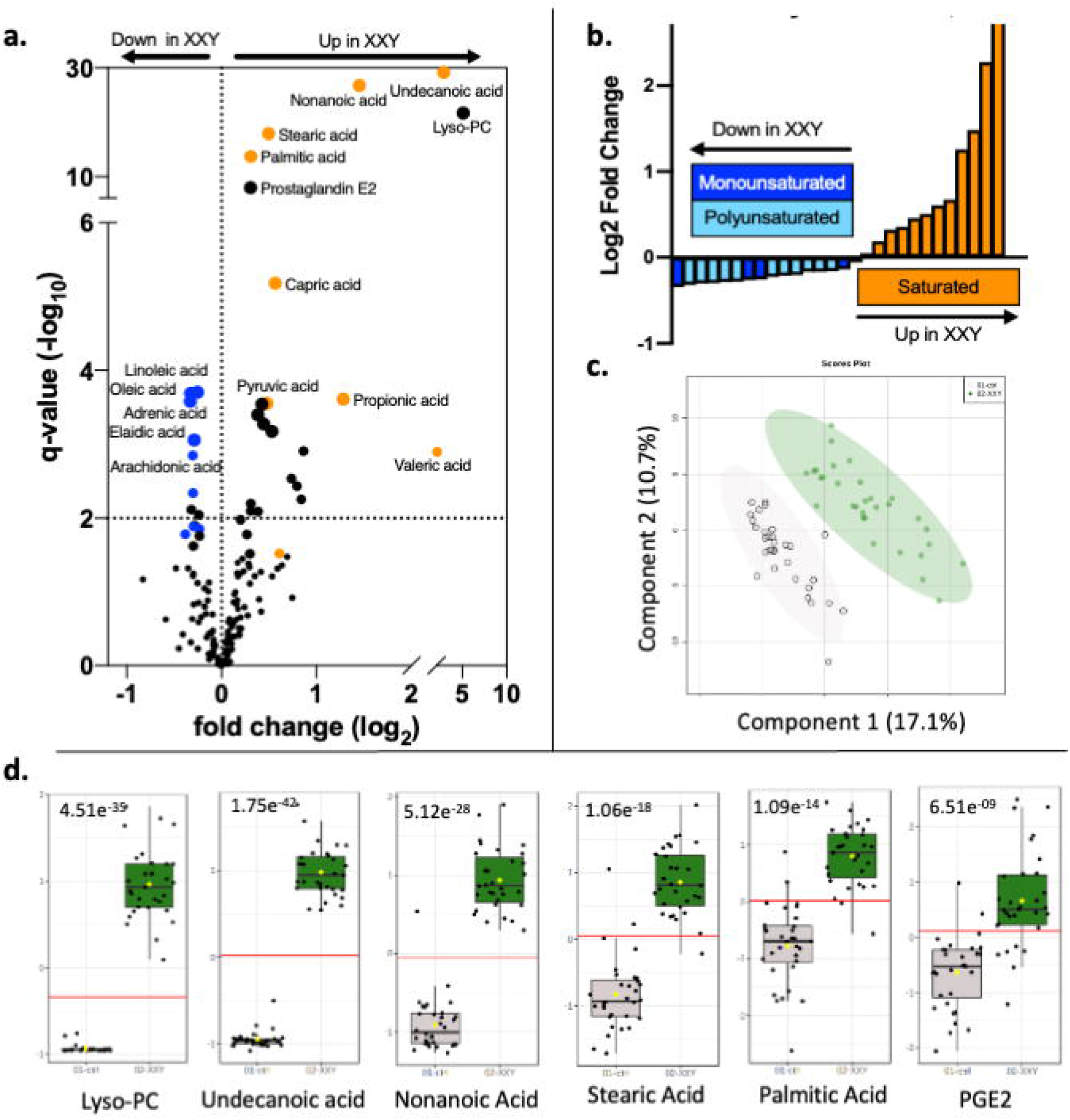
(A) Volcano plot of metabolites that are higher (right) or lower (left) in KS compared to controls. Horizontal dashed line is set at q=0.05, indicating a priori significance. Orange points represent saturated fatty acids and blue points represent unsaturated fatty acids. (B) Bar graph of fatty acids. Positive log2fold change indicates higher in KS compared to controls. (C) PLS-DA plot of the first two components, clearly differentiating KS (green) from controls (gray). (D) Individual plots of the top six metabolites loading into PLS-DA analysis as well as most significant as biomarkers of KS, with an area under the curve of >0.9 in ROC analysis. All metabolites are higher in KS (green) compared to controls (gray), with p-values in the top left corner for every metabolite. Lyso-PC=lysophosphatidylcholine. PGE2=prostaglandin E2.

**Figure 2.**
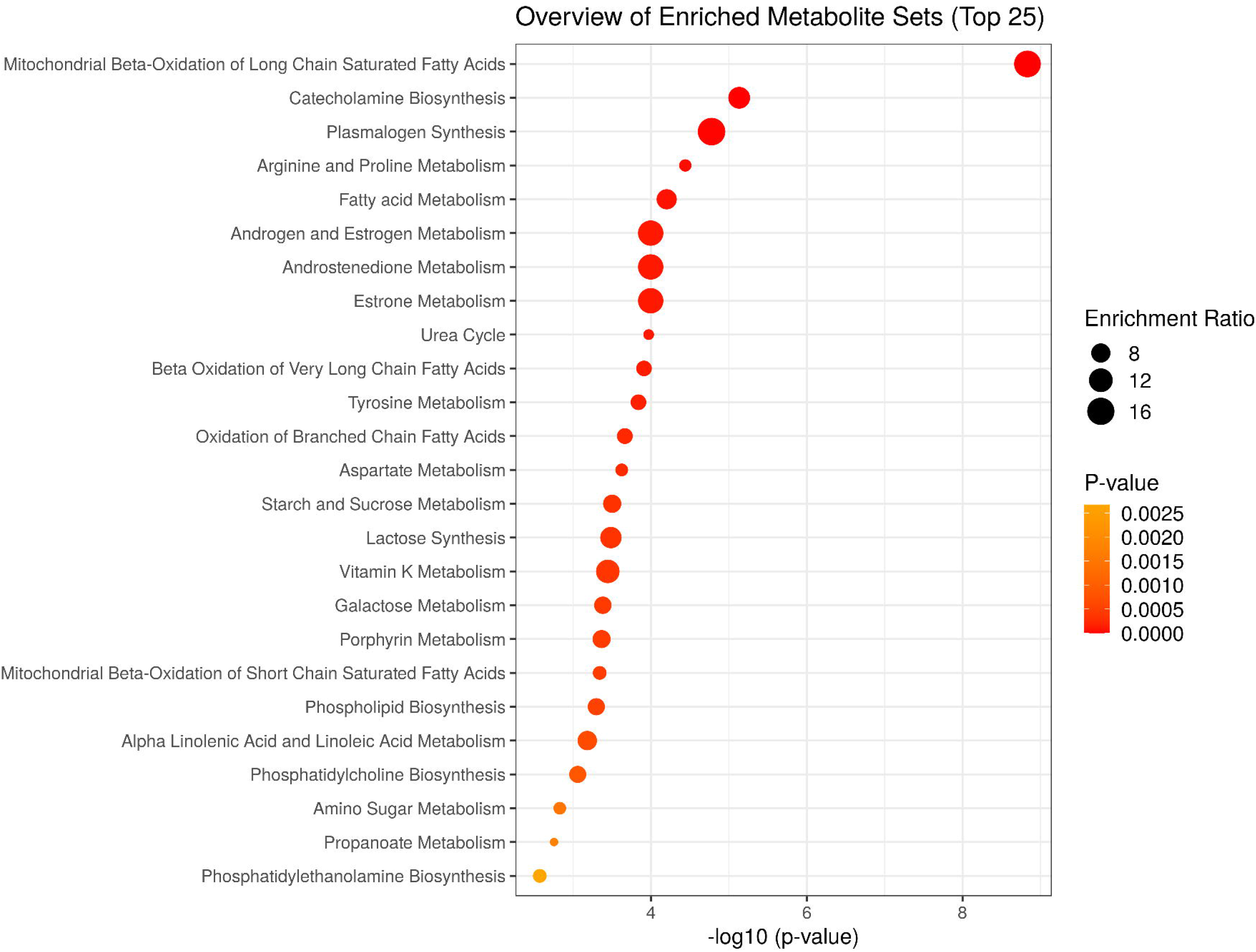
Top 25 significantly enriched metabolic sets in KS compared to controls from the SMDPB library of human metabolites. These enriched pathways suggest dysregulation of four more general ontologies of metabolism: mitochondrial, peroxisomal, amino acids, and steroid hormones.

### Testosterone-treated vs untreated

Approximately half of the KS group was treated with exogenous testosterone on a clinical basis, and there were no differences in age (p=0.94), BMI z-score (p=0.51) or systemic testosterone concentrations (380 vs 338 ng/dl, p=0.59) between treated and untreated participants. Unlike the analysis between KS and controls, no metabolites differed between testosterone-treated and untreated males with KS at an FDR of 15% (Figure 3). For exploratory purposes we proceeded to evaluate metabolites that were significantly different at a nominal p-value of <0.05. Stearidonic acid (p=0.004) and eicosapentaenoic acid (p=0.037) were lower in the testosterone-treated group, while mercaptopurine (p=0.019), threonine (p=0.024), carnitine (p=0.033), 2-oxo-7-methylthioheptanoic acid (p=0.033), lysine (p=0.041), and phenylalanine (p=0.045) were all higher in the testosterone-treated group.

**Figure 3.**
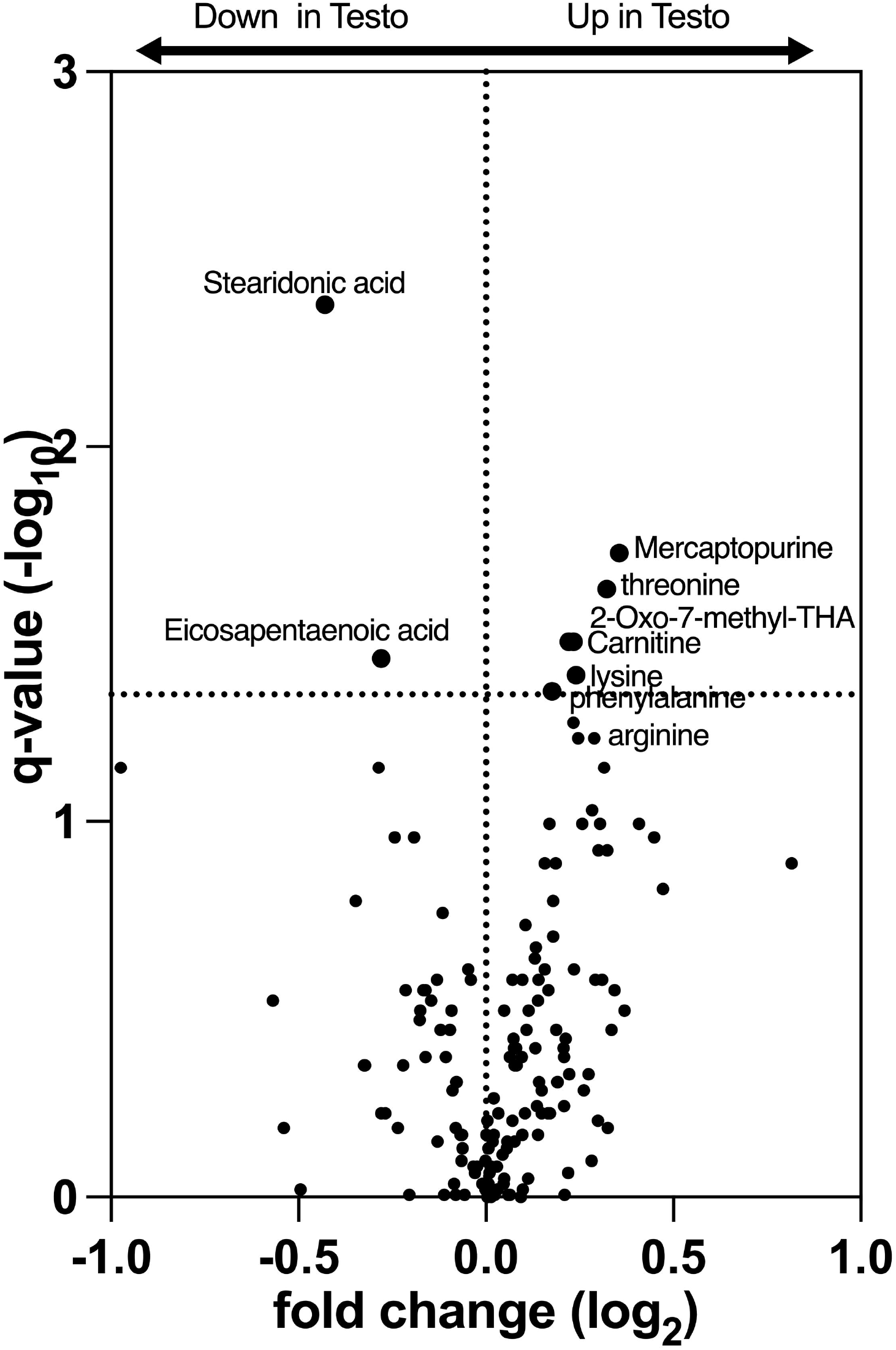
Volcano plot of metabolites that are higher (right side of X axis) or lower (left) in testosterone-treated boys with KS compared to untreated boys. The horizontal dashed line indicates a raw p=0.05 due to the lack of differences at q=0.05. Note the difference in both axes compared to Fig 1a.

## Discussion

In this exploratory study, we found a unique plasma metabolic signature in adolescent males with KS compared to male controls of similar age, BMI, and pubertal stage. The fatty acid profile was the most disparate between KS and controls, with the KS group having higher saturated fatty acids and lower unsaturated fatty acids. Mitochondrial ß-oxidation of long chain saturated fatty acids was the top enriched metabolic pathway. KS individuals on testosterone treatment were not protected from these metabolic differences, and in comparison to untreated individuals, did not result in a distinct metabolite signature. These results suggest an underlying difference in metabolism in individuals with KS that is likely independent of androgen exposure. Whether this profile is simply reflective of aberrant metabolism in KS (biomarker) or actually contributes to the pathophysiology of the KS phenotype requires further study.

Fatty acid metabolism through oxidation in the mitochondria is critical for cellular energy production. The overall metabolite pattern, both by manual analysis and pathway enrichment analysis, suggests males with KS have impaired ability to oxidize fatty acids. Unlike mitochondrial disorders that result from a single enzyme deficiency, our results suggest KS is associated with a more global impairment in mitochondrial metabolism. Many studies have identified elevated acylcarnitine esters and branched chain amino acids as markers of impaired mitochondrial function and potential contributors to insulin resistance, which was our original hypothesis for this study.^10, 20-22^ A recent study in girls with Turner syndrome found lower branched chain amino acids compared to controls, and no correlation of these metabolites with BMI z-score despite strong correlations in the control group, suggesting that obesity in Turner syndrome was not associated with altered amino acids.^23^ While we also did not observe distinct abnormalities in the metabolites typically associated with obesity and insulin resistance, the accumulation of precursors of both ß-oxidation and the tricarboxylic acid (TCA) cycle imply relative impaired mitochondrial metabolism in KS. This profile may be unique to the type of mitochondrial dysfunction in KS and these results support the need for additional investigation of in vivo mitochondrial function in males with KS, as this may be an important contributor to cardiometabolic pathology.

The finding of lower mono- and polyunsaturated fatty acid species in KS is also notable, as these are often considered “healthier” fatty acids. Kim et al found a similar profile in men with overweight/obesity compared to healthy weight.^24^ While our KS and control groups had a similar BMI, our work and that of others has shown that adiposity and other cardiometabolic risk factors are greater in KS despite normal BMI. This raises the possibility that this specific finding we observed in KS is secondary to differences in body composition among this cohort rather than a unique genetic effect from the extra X chromosome specifically. We also did not assess or standardize dietary intake in this cohort, therefore differences in diet composition may contribute to these findings, although to our knowledge participants were not taking supplements that would explain these results. Given the fatty acid profile we observed, dietary supplementation of polyunsaturated fatty acids may be a potential therapeutic avenue to further investigate in future studies in KS.

Our results yielded a few other intriguing findings beyond differences in fatty acid metabolism. First, lysophosphatidylcholine (lyso-PC) was strikingly more abundant in KS, with no overlap between groups. Lyso-PC is a normal component of blood plasma, but has been associated with metabolic and neurological disease including hyperlipidemia, atherosclerosis, ischemia, and demyelination.^25-27^ It has also been identified as a potential biomarker of obesity in men.^24^ Although this is intriguing, lyso-PC was below the limit of detection in many of our controls and thus warrants further investigation. Higher levels of the inflammatory mediator prostaglandin E2 is another finding of this study that warrants further investigation considering the cardiometabolic, neurologic, and immune system dysregulation in KS. Future directions include investigating the relationship of these disparate metabolites with the clinical phenotype.

The differences in metabolism based on testosterone treatment status was unimpressive. No metabolites were different based on our initial significance threshold, although when this was relaxed we observed that several amino acids and carnitine were higher in the testosterone-treated group, while two polyunsaturated fatty acids were lower. The largely null results were surprising given the suspected effects hormones have on metabolism. In direct comparison, red blood cell metabolites from blood donors on testosterone replacement therapy analyzed in the same laboratory used in this study revealed significant differences compared to blood from male donors not on testosterone, suggesting increased activation of antioxidant pathways and higher acylcarnitines.^28^ Our results may underestimate the impact of testosterone treatment on the metabolome due to our small sample size, significant heterogeneity in the testosterone-treated group due to diverse clinical management, and no differences in serum testosterone concentrations between treated and untreated groups (suggesting all were eugonadal either endogenously or with treatment). Nonetheless, exogenous testosterone in adolescent males with KS seems to have a minimal influence on the plasma metabolome and does not normalize the striking metabolic disparities observed between KS and controls.

To our knowledge, this is the first investigation of the plasma metabolome in individuals with KS. This study yielded several important findings for furthering our understanding of the unique metabolism in KS and speculation toward the association with clinical pathology. However, there are several important limitations. First, we utilized a convenience sample of available plasma specimens that may have introduced artificial group differences. However, we are unaware of any pre-analytical factors that would result in the profile we observed here and most of the known factors that would alter the metabolomic profile including fasting status, morning collection time, processing procedures, and lack of repeated freeze/thaw cycles were identical between groups.^29^ While the plasma metabolome has been used in many studies, it does not necessarily represent tissue-level metabolic processes. Finally, although our underlying hypothesis was that the metabolite profile would reflect differences in mitochondrial metabolism, which our results support, the approach was largely exploratory with a relatively small sample size; therefore, the inferences should be considered preliminary.

In conclusion, the plasma metabolomic profile in adolescent males with KS is distinctly different from their peers of similar BMI and pubertal stage. This presents an opportunity to further our understanding of the underlying molecular pathology involved in KS, particularly with regard to mitochondrial metabolism.

## Data Availability

Data will be provided upon reasonable request.

## Acknowledgements

The authors would like to thank the individuals who participated in this study. Funding support included Pediatric Endocrine Society Career Scholar Award, Endocrine Fellows Foundation Junior Faculty Award, Child Maternal Health CCTSI Pilot Award (UL1TR002535), and the NICHD K23HD092588. Contents are the authors’ sole responsibility and do not necessarily represent official NIH views.

## Notes

**Disclosure summary:** No authors have conflicts of interest to report

### Competing Interest Statement

The authors have declared no competing interest.

### Clinical Trial

NCT02723305

### Funding Statement

Pediatric Endocrine Society Clinical Scholars Award; National Institute of Child Health and Human Development (NIH/NICHD) K23HD092588; 
National Center for Advancing Translational Sciences (NIH/NCATS) Colorado CTSA UL1 TR002535. 

### Author Declarations

The study was approved by the Colorado Multiple Institutional Review Board (COMIRB). Written informed consent was obtained prior to any study procedures.

